# Study protocol to stablish a prospective cohort for the study of phenotypic clusters, progression paths and outcomes of frailty and dependence: The CohorFES

**DOI:** 10.1101/2025.05.29.25328600

**Authors:** Natàlia Garcia-Giralt, Diana Ovejero, Jose Antonio Carnicero Carreño, Anna Ribes, Pedro Abizanda Soler, Jose Antonio Serra Rexach, Francisco José García García, Montse Rabassa, Leocadio Rodriguez Mañas, Àlex Sanchez Pla, Mariam El Assar de la Fuente, Carmen Maria Osuna Del pozo, Inmaculada Carmona, María Ángeles Caballero-Mora, Virginia Mazoteras Muñoz, Elisa Belen Cortes Zamora, Almudena Avendaño Céspedes, Bárbara Agud Andreu, Fernando Gómez Galera, Jade Soldado-Folgado, Maria Cristina Andrés Lacueva, Xavier Nogués

**Affiliations:** CIBER on Frailty and Healthy Ageing (CIBERFES), ISCIII, Spain; Hospital del Mar Research Institute, Barcelona, Spain; Fundación para la Investigación Biomédica del Hospital Universitario de Getafe. Instituto de Investigación Sanitaria Hospital Universitario de Getafe (IISGetafe), Spain; Complejo Hospitalario Universitario de Albacete, SESCAM, Spain; Hospital General Universitario Gregorio Marañón Madrid, Spain; Complejo Hospitalario de Toledo · SESCAM, Spain; Departament de Nutrició, Ciències de l’Alimentació i Gastronomia, Nutrition and Food Safety Research Institute (INSA), Facultat de Farmàcia i Ciències de l’Alimentació, Universitat de Barcelona (UB), Spain; Departament de Genètica, Microbiología i Estadística. Universitat de Barcelona, Spain; Servicio de Geriatría, Hospital General Universitario de Ciudad Real, SESCAM, Ciudad Real and Instituto de Investigación Sanitaria de Castilla la Mancha, IDISCAM, Spain; Laboratori de Referència de Catalunya. Hospital del Mar. Barcelona, Spain

**Author notes:** Corresponding author: Natalia Garcia-Giralt, PhD, CIBER on Frailty and Healthy Ageing, Hospital del Mar Research Institute, Department of Genetics, Microbiology and Statistics, University of Barcelona.

**Keywords:** Frailty, CohorFES, FTS5, metabolomics

## Abstract

Frailty has become a major problem for the health system, but also a window of opportunity to fight against disability through preventive strategies focused on the detection and treatment of frailty in all settings. However, no systematic strategies of screening and early detection are available in clinical settings. This project aims to identify clinical and biological phenotypic clusters that drive through the different stages of frailty and to describe the underlying mechanisms of the trajectories leading to disability and the potential for treatment. Moreover, validation of Frailty Trait Scale 5 (FTS5) will be performed as an easy model to be implemented in primary care and hospital scope.

A prospective population-based cohort will be stablished for frailty phenotyping (CohorFES). Creation of a CIBERFES Biobank where blood and urine samples from participants of CohortFES are stored for future researches. Demographic and clinical history data, anthropometric measurements, predimed questionnaire, peripheral blood biochemical variables, and metabolomics were collected for each participant at baseline and every year until become frailty.

Using cluster partition models (k-means and hierarchical clustering) will group together individuals with similar deficits and characteristics (frailty phenotypes). Then, by using pre-established criteria (gap and silhouette), the proposed clustering solution (belonging to given clusters) will be evaluated. Further, we will assess, in a longitudinal fashion, the appearance and accumulation of deficits during the study period and identifying the clusters subgroups with more rapid progression. Results will be applied to establish and compare clusters and trajectories. Finally, frailty phenotypes and patient clusters will be correlated with health outcomes such as the use of health services (both primary and secondary care), hospital admissions, and mortality.

Information about clinical and biological phenotypic clusters that drive through the different stages of frailty can lead to identify potential targets that could improve the therapeutic management of these patients.

In summary, from a research perspective the project aims to better understanding of the interindividual variability in clinical events that lead to frailty, dependence and finally, to death.

**Protocol Registration:** NCT06965972 (date 05/02/2025)

## 1. Introduction

Frailty is one of the major challenges of the 21st Century, and a top priority for national and international organisms like the WHO (World Health Organization) or the European Parliament. This has put frailty as one of the top priorities in the biomedical research agenda of the European Commission [1,2]. Frailty is constituted by a physiological state of increased vulnerability and impaired resilience to stressors (i.e. diseases, external agents, drugs tolerability and toxicity) due to the combined effect of the aging process and some chronic diseases which drives to a final stage of dependency and disability with a sharp impact in quality of life, health and social resources consumption, hospitalization and death [2].

It is well-known the relevance of frailty, its detection, and management since we are aware about their reversibility [3], the costs on the health systems [4], and its potential impact in clinical settings [5]. In a clear contrast with the abundancy of data in non-clinical settings, there is a lack of strong data in the clinical setting where the prevalence of frailty is higher and where the risks for developing its most serious adverse consequences is more likely [6]. There is hence an urgent need for a better screening and diagnosis of frailty, its trajectories and the determinants of these separate trajectories depending upon both the characteristics of frailty in each patient (associated or not with sarcopenia, or cognitive impairment or different clusters of chronic diseases).

### Review of prior research

While the different categories of the syndrome based on the severity of the observed deficits (robust, frail, pre-frail) are quite well defined and characterized from an epidemiological point of view [7,8], there is a scarcity of data on the functional pathways between these diagnostic categories (and, among them, disability), and this is especially true in clinical cohorts. This is really shocking considering that one of the most relevant factors, if not the first one, associated with a poor evolution of frailty is to experience an episode of hospitalization [9].

Another important issue to address is to implement an easy tool to identify frailty and related factors in our full outpatients list. The Frailty Trait Scale-FTS is able to detect frailty in some clinical settings (Acute Care Geriatric Unit, Geriatric Service outpatient office and Primary Care), with a good predictive capacity for adverse outcomes (death, incident disability, deterioration in SPPB, falls and hospitalization) at 6-12-18 months [10,11]. However, the full version of FTS, composed of 12 items, takes around 15 minutes, making it unpractical in usual clinical conditions, where the time available by the physician or the nurse is lower. With this fact in mind, a shorter version of only 5 items (the so-called FTS 5) was developed [12]. This shorter version offers promising results based upon the sensitivity to detect small changes shown by the full FTS. Finally, the variables that compose the FTS5 (gait velocity, grip strength, BMI, PASE, and balance) can be incorporated into electronic instruments as for example the electronic frailty index (eFI) [13]. The use of easy electronic tools has been useful not only in hospital care but also in routine primary care practice. Moreover, it would be easier to measure the adverse outcomes, including falls, delirium, disability, care home admission, hospitalization and mortality as it has been recently shown [14].

In this line, several clinical guides have been proposed for early identification and prevention of frailty in elderly people [15, 16, 17]. These guides try to address one of the major health challenges such as the increase in ageing populations and the growing incidence of chronic illnesses. In particular, in 2022, the Frailty and Falls Prevention Working Group updated a “consensus document on prevention of frailty in elderly people” approved by the Interterritorial Council of the Spanish National Health System. This document is focused on the importance of early diagnosis and the need for interventions within the health care system, especially in primary healthcare [15]. It is important since frailty is not inherent to the ageing process itself, but that it is potentially reversible, even spontaneously, especially in the early stages. Early detection and diagnosis of frailty is therefore essential in terms of life quality, but also in terms of the use of health and social resources.

Our overarching goal is therefore to identify the critical subgroups of subjects at risk of progression from robustness to prefrailty and frailty and from there to their late stages, and the pathways that mediate this trajectory amongst community-dwelling Spanish subjects.

## 2. Project objectives

Inside this conceptual framework and considering the scarce data available in clinical settings about frailty diagnosis, trajectories and prognosis, the main goal of this project is to stablish a clinical, real-life and prospective cohort (COHORFES) to identify clinical and biological phenotypic clusters that drive through the different stages of frailty and to identify the underlying mechanisms that finally will trigger the disability. Moreover, validation of Frailty Trait Scale 5 (FTS5) will be performed as an easy model to be implemented in Primary care and Hospital scope.

Finally, the creation of a multicentered biobank inside the CIBERFES consortium where blood and urine samples from participants of the COHORFES are stored for future researches.

## 3. Methodology and implementation

### 3.1 Study design and setting

We have stablished a multicentered cohort, the COHORFES, **a prospective and observational study** based on population. The study is registered in clinicaltrials.Gov as observational study: NCT06965972 (date 05/02/2025). Patients are recruited from the beginning of the project and followed year on year during all the study time. Participants are women and men 65 years old or above visited in the outpatient clinics of participant centers:

Hospital del Mar Research Institute, Barcelona Hospital General Universitario Gregorio Marañón, Madrid Complejo Hospitalario Universitario de Albacete, Albacete Hospital General Universitario de Ciudad Real, Ciudad Real Fundación del Hospital Nacional de Parapléjicos, Toledo Hospital Universitario de Getafe, Madrid

### 3.2 Participant selection for cohort study

Inclusion criteria

- women and men >65 years old
- Signed written informed consent

Exclusion criteria

Patients in a critical situation of end of live or Barthel scale <60. Pacients with a hospitalization for any disease in the last 6 months Pacients whith diagnosis and current treatment of any type of cancer except cutaneous no melanoma

### 3.3 Sampling procedure

Individuals visited in participant centers and meet inclusion criteria are asked to participate into the study. These individuals are consecutively included to the study after signed the informed consent. All individuals of the COHORFES are invited to participate in the BioBank after informed consent was obtained. Serum, plasma, buffy coat, and urine samples will be collected at baseline (V0) and every year after their cohort inclusion.

### 3.4 Sample size

The calculation of the sample size in this study is based on index data obtained from the Study of Toledo of Healthy Aging. As is expected that frailty prevalence rates as well as the progression of the frailty syndrome will be superior in the clinical population that in the general population, these estimates can be considered as conservative and they allow, therefore, an additional margin of security in reaching sufficient statistical power. Frailty rates are estimated from subjects who have a negative FTS5 that later develop frailty, which was 0.075 (7.5%). The ratio of exposed / unexposed was 1008/196 (ratio of negatives vs. positives), that is 5.1. The minimum relative risk (RR) to be detected is estimated taking as an interest variable “incident frailty” (stage transition marker) for which OR of the different estimates range between 1.78 and 5.4 and therefore we adopt a RR conservative to detect of 2. Finally, the loss of follow-up rates is estimated at 10%. With these data, accepting an alpha risk of 0.05 and a beta risk of 0.2 in a bilateral contrast, 215 subjects in the exposed group and 1075 in the unexposed group are needed to detect a factor of risk with a minimum RR of 2. The POISSON approach has been used for this calculation.

### 3.5 Study schedule

This is a prospective study with long-term follow-up where patients will have a visit every 12 months for all procedures. We anticipate follow-up until 1500 patients are included with a minimum follow-up of one year. The recruitment period comprises from January 2, 2021, to December 31, 2028. Data will be collected during all the study period which is estimated to finalize at 2030. At this time, all data will be analyzed and finally reported.

Each participant will perform minimum 3 visits. The study visits will be the following:

- A preselection visit to check inclusion/exclusion criteria and to inform to the participant about the project.
- A start visit at baseline (V0) to obtain all the study’s variables and the informed consent of the participant
- A follow-up visit each 12 month until end of the study. Same clinical variables than baseline are collected. Samples are also collected.
- A final study visit:
  a. Final of the project
  b. Participant withdraws
  c. Frailty diagnosis or dead

### 3.6 Variables

All variables and biological samples will be obtained at baseline and in all the successive visits until the end of the study or the patient withdraw.

#### Main variables

The main variables are based on the evaluation of the frailty score:

- Frailty Trait Scale 5 ítems: 1.-walking speed test, 2.-grip strength, 3.-Physical Activity, 4-Body Mass Index (BMI), 5.-progressive Romberg test.
- Fried phenotype
- Electronic Frailty index

#### Secondary variables

For each patient, the following data is obtained from the Clinical History and by questionnaires:

1. *Demographic data:*
  - Age in years and date of birth
  - Sex
  - Living situation
  - Educational level
2. *Clinical history data*
  - Prevalent diagnosis
  - Loss of weight in the last year
  - Geriatric Depression Scale: The Geriatric Depression Scale (GDS) is a self-report measure of depression in older adults. Users respond in a “Yes/No” format. The GDS was originally developed as a 30-item instrument. Since this version proved both time-consuming and difficult for some patients to complete, a 15-item version was developed. The shortened form (GDS-S) is comprised of 15 items chosen from the Geriatric Depression Scale-Long Form (GDS-L). These 15 items were chosen because of their high correlation with depressive symptoms.
  - Barthel Index: Barthel Index is an ordinal scale used to measure performance in basic activities of daily living i.e. feeding, bathing, grooming, dressing, bowel control, bladder control, toileting, chair transfer, ambulation and stair climbing. The index also indicates the need for assistance in care. It is a widely used measure of functional disability. The index was developed for use in rehabilitation patients with stroke and other neuromuscular or musculoskeletal disorders, but may also be used for oncology patients.
  - Lawton-Brody Instrumental Activities of Daily Living Scale: is an instrument to assess independent living skills. It takes approximately 10 to 15 minutes to administer and contains 8 items that are rated with a summary score from 0 (low functioning) to 8 (high functioning).
  - Pfeiffer test: Also known as Short Portable Mental Status Questionnaire (SPMSQ), it is a brief test of 10 questions used to assess a person’s cognitive status, especially in older adults. It evaluates aspects such as memory, temporal and spatial orientation, and the ability to perform basic tasks.
  - Predimed questionnaire: The adherence of participants to the Mediterranean diet will be assessed through the 14-item Mediterranean diet adherence screener (MEDAS) validated for the Spanish population in a phone interview with the participant [20]. It consists of two questions about eating habits, eight questions about the frequency consumption of typical foods of the Mediterranean diet, and four questions about the consumption of foods not recommended in this diet. Each question is scored with 0 (non-compliant) or 1 (compliant), and the total score (from 14 questions) ranged from 0 to 14, so a score of 14 points means maximum adherence [20].
3. *Anthropometric measurements* (in the case of the center has the facilities and densitometer): Total body Dual-Energy X-Ray Analysis by using a DXA device for the measurement of Bone Mineral Density (both overall and in the usual regions of interest) and body composition (fat mass, muscle mass).
4. *Peripheral blood biochemical variables*
  - Leukocytes ml/mmc
  - Red blood cells ml/mmc
  - Hemoglobin g/dl
  - Hematocrit %
  - Red blood cell distribution width %
  - Platelets ml/mmc
  - Vitamin D ng/ml
  - TSH nmol/l
  - Glucose mg/dl
  - Glycated hemoglobin (diabetics only) %
  - Creatinine mg/dl
  - Sodium meq/dl
  - Potassium meq/dl
  - Calcium mg/dl
  - Phosphorus mg/dl
  - GPT u/l
  - GOT u/l
  - Phosphatase alkaline u/l
  - Total proteins g/dl
  - Albumin g/dl
  - Prealbumin mg/dl
  - Cholesterol mg/dl
  - Triglycerides mg/dl
  - HDL mg/dl
  - LDL mg/dl
  - C-reactive protein mg/ml
5. *Metabolomic analysis*
  a. Exposome assay A targeted quantitative metabolomics approach will be applied to analyze the plasma samples using liquid chromatography coupled with tandem mass spectrometry (LC-MS/MS), as described previously [18, 19). This method will employ analyte extraction and LC separation combined with selective mass-spectrometric detection using multiple reaction monitoring (MRM) pairs to identify and quantify metabolites. The approach will allow for the comprehensive profiling of over 1100 metabolites, including dietary-derived metabolites (especially (poly)phenols and their microbiota-derived derivatives); other dietary markers such as non-(poly)phenolic compounds (e.g., glucosinolates, methylzanthines, alkaloids, amino acid derivatives such as peptides, S-allylcysteine, fatty acids, benzoxazinoids); environmental and lifestyle-related metbolites (such as pollutants, pharmaceuticals, nicotine and alcohol metabolites); and approximately 500 endogenous compounds involved in key metabolic pathways (e.g., amino acids and derivatives, lipids, vitamins, biogenic amines, organic acids, and carbohydrates). The panel will also include metabolites from the tryptophan-kynurenine pathway, as well as and microbial-derived metabolites such as indoles, bile acids, and short-chain fatty acids and others. Plasma samples will be stored at −80 °C until analysis. For extraction, 10 μL of internal standard solution will be added to 100 μL of plasma, followed by 500 μL of cold acetonitrile (−20 °C) containing 1.5 M formic acid and 10 mM ammonium formate in the plate. After vortexing and incubation at −20 °C to induce protein precipitation, the extracts will be collected using a Waters Positive Pressure-96 processor (Waters, Milford, MA, USA), dried under a stream of nitrogen gas, and reconstituted in 100 μL of water:acetonitrile (80:20, v/v) containing 0.1% formic acid (v/v) and external standards. The samples will then be vortexed and prepared for injection. Mass spectrometric analysis will be performed on a Sciex 7500 QTrap® instrument coupled with an Agilent 1290 UHPLC system. A Luna Omega Polar C18 column will be used for chromatographic separation, with distinct different gradients applied for positive and negative ion modes. The instrument will operate in scheduled MRM (sMRM) mode to ensure high sensitivity and selectivity for all targeted metabolites.
  b. Clinical biomarkers assay Plasma samples will be prepared and analysed using a combination of direct flow injection tandem mass spectrometry (DFI-MS/MS) and liquid chromatography tandem mass spectrometry (LC-MS/MS) (20). This targeted clinical biomarker assay will aim to provide a robust and user-friendly platform for the discovery and quantification of metabolomic biomarkers. The method will allow the targeted identification and quantification of up to 651 metabolites from 18 different analyte groups, including amino acids and derivatives, biogenic amines, organic acids, nucleotide/nucleosides, lipids, acylcarnitines, and glycerophospholipids. The analytical workflow will combineanalyte derivatization, extraction and high-throughput quantification using MRM in tandem mass spectrometry. For accurate quantification, the method will incorporate stable isotope-labelled internal standards, along with other compound-specific internal standards, across all metabolite groups.
6. *Healthcare resource use:*
  - Number of Primary Care consultations
  - Number of prescribed drugs (polypharmacy)
  - Number of contacts with the Secondary Care facilities during the study period.
  - Non-elective hospital admissions. Number of admissions and length of stay during the study period.
7. *All-cause mortality*.

### 3.7 Statistical analysis plan

The cluster partition models (k-means and hierarchical clustering) will group together individuals with similar deficits and characteristics (frailty phenotypes). Then, by using pre-established criteria (gap and silhouette), the proposed clustering solution (belonging to given clusters) will be evaluated.

In a second step we will assess, in a longitudinal fashion, the onset and accumulation of deficits during the study period and identifying the clusters subgroups with more rapid progression. Results will be applied to establish and compare clusters and trajectories.

Finally, frailty phenotypes and patient clusters will be correlated with health outcomes such as the use of health services (both primary and secondary care), hospital admissions, and mortality.

## 4 Data collection and management

We use a data collection platform (REDCAP) designed by the Hospital Universitario de Getafe accessible for all project researchers who are including participants. Data from REDCAP can be exported to several applications (excel, SPSS, …) by the PI of the project. Anonymized data will be managed by coordinator researchers for the project analyses.

## 5 Potential bias

All the centers involved in the participant recruitment have specific outpatient clinic focused on frailty and older people management. These centers have a disperse geographic distribution around Spain which decreases the potential bias.

## 6 Limitations

The main limitation of the project is the coordination of patient recruitment among all participant centers since the data homogeneity and sample size is a key factor for the establishment of the COHORFES.

## 7 Ethical consideration

The study follows the national and international standards (Declaration of Helsinki and Tokyo) on ethical aspects. The protocol of this study was approved by the CEIm Parc de Salut Mar (CEIC : 2019/8622/I). This study respects the Code of Good Scientific Practices (http://www.imim.es/imim/cas/c-CBPC.htm).

Data confidentiality: the data included is pseudo-anonymized and identified by an internal code that is created at the time of its entry into the REDCAP database, which makes it impossible to identify the subjects included. Only clinical researchers of the study have access to the identifying data of the patients. The database will only contain the internal code. Project data will be stored on REDCAP at the institution.

The responsibility for the global file corresponds to the Hospital Mar Research Institute (IMIM) in Barcelona. The IMIM and the Principal Investigator of the project are responsible for the processing of the data in the framework of this study. No copies will be made outside the REDCAP server.

In all the steps of the study, the confidentiality of the included subjects is guaranteed, in accordance with the provisions of Organic Law 3/2018, of December 5, on the protection of personal data and the guarantee of digital rights (LOPD 3 /2018).

The study is subject to the ethical standards of biomedical research in humans. Patients are informed and given an informed consent document approved by the local ethics committee. Aspects related to confidentiality follows established regulations. Participants are informed, as provided for in Organic Law 3/2018, that the data may be subject to automated processing and of the rights that participants have to consult, modify or delete the personal data file. They are reported that the database is manipulated by the principal investigator and project researchers using an access key and the name and clinical history of the patient will not appear in the record.

## 8 Discussion

Frailty is and will be one of the major challenges of our society due to the increase of ageing people and its implications for health systems and social care policy [15, 16, 17]. Although in the recent years the knowledge about the management of this condition has increased, little is still known on the events and mechanisms involved in the way from robust to frailty and dependence.

Scientifically, the current project can provide valid information about clinical and biological phenotypic clusters that drive through the different stages of frailty. Moreover, we aim to identify the biological markers involved in the frailty progress for an early identification and monitoring the rapid deterioration of some patients. Therefore, identifying these pathways can lead to identify potential targets that could improve the therapeutic management of these patients.

The proposal offers:

1. An early detection of individual patients at risk of progression to frailty and dependence when fulfilling one of the identified phenotype profiles.
2. Monitoring the progression over time in the degree of frailty and vulnerability in the individual patient following an identified path of risk of evolution.
3. Offer standardized, well-defined populations where interventions to prevent or reverse the progression of frailty should be targeted.
4. Provide information for a correct identification of homogeneous clusters suitable for clinical and epidemiological research.
5. Open future research on biological markers of frailty and vulnerability progression.

The overarching goal is to minimize the impact of frailty and its consequence, dependence, improving the diagnostic classification, prognostic evaluation and management strategies in different phenotypes of risk.

## Data Availability

No datasets were generated or analysed during the current study. All relevant data from this study will be made available upon study completion.

## Funding

The present study is funded by CIBER on Frailty and Healthy Ageing, by Francisco Soria Melguizo Fundation, and FIS (ISCIII) num. PI19/00033. Additional support was provided by AGAUR (2021 SGR 00043), and FEDER, EU.

## Authors Contributions

Conceptualization: Xavier Nogués,, Jose Antonio Carnicero Carreño, Leocadio Rodriguez Mañas.

Supervision: Xavier Nogués,, Jose Antonio Carnicero Carreño, Leocadio Rodriguez

Mañas, Pedro Abizanda Soler, Jose Antonio Serra Rexach, Francisco José

García García, and Maria Cristina Andrés Lacueva

Methodology and Sample Collection: Natàlia Garcia-Giralt, Diana Ovejero, Anna Ribes,

Mariam El Assar de la Fuente, Carmen Maria Osuna Del pozo^5^, Inmaculada Carmona, María Ángeles Caballero-Mora, Virginia Mazoteras Muñoz, Elisa Belen Cortes Zamora,

Almudena Avendaño Céspedes, Bárbara Agud Andreu, Fernando Gómez Galera, and Jade Soldado-Folgado.

Validation: Xavier Nogues, Natalia Garcia-Giralt, Montse Rabassa, and Àlex Sanchez Pla.

Funding acquisition: Xavier Nogues and Natalia Garcia-Giralt

Writing – original draft: Natalia Garcia-Giralt and Montse Rabassa

Writing – review & editing: All authors

## Notes

### Competing Interest Statement

The authors have declared no competing interest.

### Author Declarations

this protocol was approved by the CEIm Parc de Salut Mar (CEIC : 2019/8622/I).

